# Solid fuel use in relation to dementia risk in middle-aged and older adults: A prospective cohort study

**DOI:** 10.1101/2022.05.03.22274530

**Authors:** Fan Pu, Yingying Hu, Chenxi Li, Xingqi Cao, Zhenqing Yang, Yi Liu, Jingyun Zhang, Xueqin Li, Yongli Yang, Wei Wang, Xiaoting Liu, Kejia Hu, Yanan Ma, Zuyun Liu

**Author notes:** **Corresponding author:** Zuyun Liu, Professor, Department of Big Data in Health Science School of Public Health and Center for Clinical Big Data and Analytics of the Second Affiliated Hospital, Zhejiang University School of Medicine, 866 Yuhangtang Rd, Hangzhou, 310058, Zhejiang, China., or. These authors contributed equally to this work (Fan Pu, Yingying Hu, Chenxi Li). These authors contributed equally as the senior author (Kejia Hu, Yanan Ma, Zuyun Liu).

## Abstract

**Objectives:** It remains unknown whether household air pollution is associated with dementia risk. This study examined the associations between solid fuel use for cooking and heating (the main source of household air pollution) and dementia risk.

**Methods:** This analysis included data on 11,352 participants (aged 45+ years) from the 2011 wave of China Health and Retirement Longitudinal Study, with follow-up to 2018. Dementia risk was assessed by a risk score using the Rotterdam Study Basic Dementia Risk Model (BDRM) and then standardized for analysis. Household fuel types of cooking and heating were categorized as solid (e.g., coal, crop residue) and clean (e.g., central heating, solar). Multivariable analyses were performed using generalized estimating equations. Moreover, we examined the joint associations of solid fuel use for cooking and heating with the BDRM score.

**Results:** We found an independent and significant association of solid (vs. clean) fuel use for cooking and heating with a higher BDRM score after adjusting for potential confounders (e.g., ß = 0.14 for solid fuel for cooking; 95% CI: 0.12, 0.17). Participants who used solid (vs. clean) fuel for both cooking and heating had the highest BDRM score (ß = 0.20; 95%CI: 0.16, 0.23). Subgroup analysis suggested stronger associations in participants living in rural areas.

**Conclusions:** Solid fuel use for cooking and heating was independently associated with increased dementia risk in Chinese middle-aged and older adults, particularly among those living in rural areas. Our findings call for more efforts to facilitate universal access to clean energy for dementia prevention.

## 1. Introduction

Dementia currently affects more than 50 million people worldwide, with the number expected to reach 152 million by 2050. In the absence of definitive and effective treatment for dementia, early prevention is a very important strategy for reducing the dementia risk. This is an important issue especially for low- and middle-income countries (LMICs), which have more patients (i.e., nearly two-thirds of global patients) but fewer dementia-related medicare resources compared to high-income countries (HICs). The identification and modification of risk factors remain fundamental to developing primary and secondary prevention strategies, to lower the incidence of dementia.

Prior studies have shown that ambient air pollutants, including CO, PM2.5, NOx, and O_3_, are associated with increased dementia risk (Chen et al. 2017; Jung et al. 2015; Li et al. 2019; Oudin et al. 2016). However, in modern society, people spend 90% of their lives indoors. Household air pollution (HAP) has been established to have a more significant effect on health than outdoor air pollution (Apte and Salvi 2016; Kurmi et al. 2012; Tran et al. 2020). As the main contributor to HAP, solid fuel use for cooking and heating causes 4 million premature death per year globally (Daly and Walton 2017). In China, solid fuels are the primary source of household energy for over 700 million people (Banerjee et al. 2012; Smith et al. 2013). Moreover, household solid fuel use has been linked to a variety of diseases, including cataracts (Ravilla Thulasiraj et al. 2016), tuberculosis (Kolappan and Subramani 2009), and cardiovascular disease (CVD) (Painschab et al. 2013). Dementia is highly prevalent in an aging society. Nonetheless, the potential effects of HAP from the use of solid fuel on dementia have not been evaluated yet.

To the end, we leverage survey data from China Health and Retirement Longitudinal Study (CHARLS), a large and prospective nationally representative cohort study of middle-aged and older adults, who are the most sensitive age groups to both air pollution and dementia. The present study aimed to demonstrate the associations of solid fuel use for cooking and heating with dementia risk, assessed using a risk score based on the widely used Rotterdam Study Basic Dementia Risk Model (BDRM) (Licher et al. 2019). We also evaluated whether the associations differed by important socialdemographic characteristics such as age, sex, and residence.

## 2. Methods

### 2.1. Study design and participants

This study used data from the 2011-2012, 2013-2014, 2015-2016, and 2018-2019 waves of the CHARLS. CHARLS is a national longitudinal cohort study covering 450 urban communities and rural areas in 28 provinces of China. The survey includes participants’ information on household demographics, health status, health care utilization, insurance, and other variables. Data can be accessed through its official website (charls.ccer.edu.cn/en). Additional details on study design regarding sampling, response rates, and data quality assessment have been published previously (Zhao et al. 2014). CHARLS was approved by Peking University’s Ethical Review Committee (IRB000001052-11015). All participants provided written informed consent prior to participation.

The baseline survey was launched from 2011 to 2012 and recruited 17,708 participants. We excluded participants whose ages were under 45 years old (N = 484), with missing data on demographics (N = 1,569) and household energy source (N = 204); then, we excluded participants with missing data on variables used to calculate the BDRM score in 2011 wave (N = 192) and in all follow-up waves (N = 3907), leaving 11,352 participants for the final analysis. Details of the flowchart are shown in Figure 1.

**Fig. 1.**
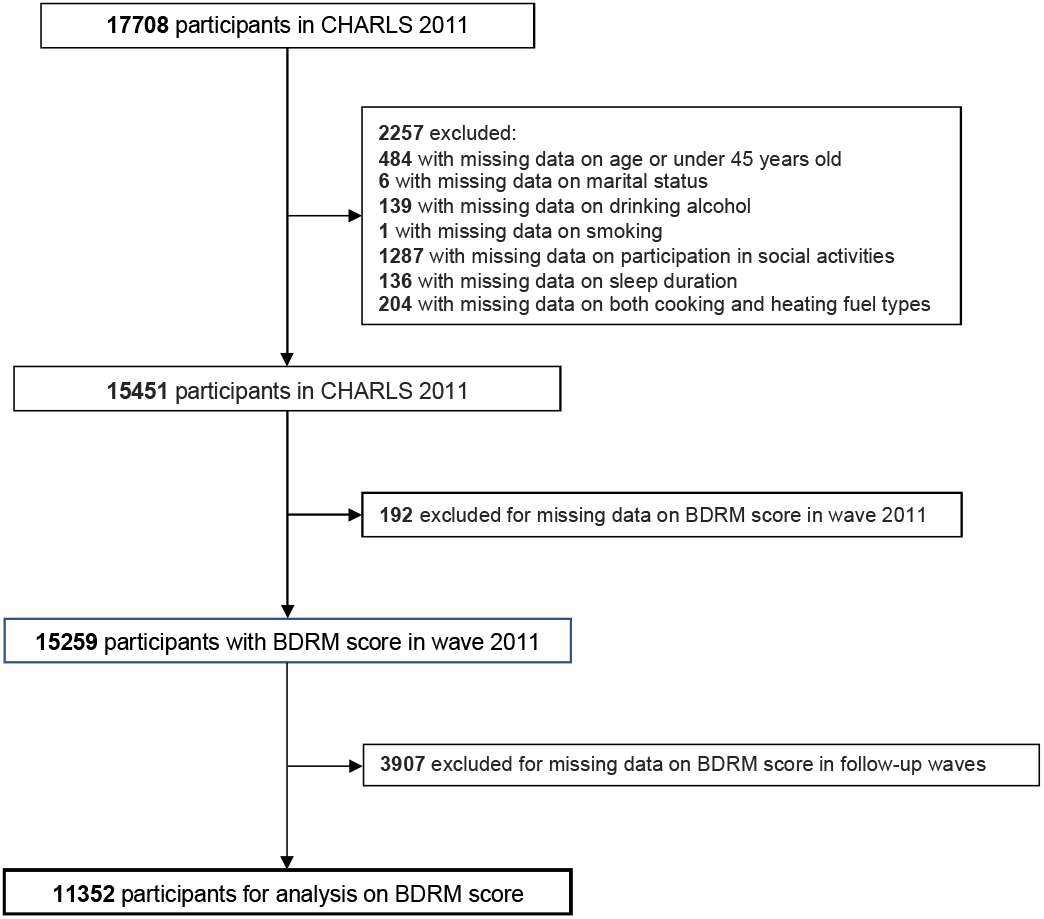
Flow chart of the selection process of participants.

### 2.2. Dementia risk measurement

Dementia risk was measured at the baseline and in follow-up waves using the BDRM, a dementia risk prediction model developed in HICs and showed similar discriminative ability when applied in LMICs (Stephan et al. 2020). The BDRM score was quantified by weighing the following risk factors to predict dementia: age (1.09 points), history of stroke (1.82 points), subjective memory decline (1.31 points), and need for assistance with finances or medication (1.46 points). The detailed scoring algorithm is shown in Table S1. BDRM scores were first calculated and then z-score standardized for the statistical analysis, with higher scores indicating greater dementia risk.

### 2.3. Household fuel types

Household fuel types of cooking and heating were assessed by a structured questionnaire with the following questions: “What is the main source of cooking fuel? (1) Coal, (2) Natural gas, (3) Marsh gas, (4) Liquefied petroleum gas, (5) electric, (6) Crop residue/Woodburning, (7) Other”, and “Does your residence have heating? (1) Yes, (2) No’ and ‘What is the main heating energy source? (1) Solar, (2) Coal, (3) Natural gas, (4) Liquefied petroleum gas, (5) electric, (6) Crop residue/Woodburning, (7) Other”. Household fuel types of cooking and heating were categorized as solid (i.e., coal, crop residue, wood) and clean (i.e., central heating, solar, natural gas, liquefied petroleum gas, electric, or marsh gas). We excluded participants whose answer was “other” as we could not precisely assess the type of fuel.

### 2.4. Covariates

Covariates including sociodemographic factors and health behaviors were obtained at baseline. The sociodemographic variables including sex, educational level, marital status, residence, and social activities were collected from the self-reported questionnaire. Educational level was categorized into lower than middle school or middle school and above. Marital status was categorized into currently married or others (e.g., separated, divorced, widowed). The residence was categorized into rural and urban. Individuals were considered to participate in social activities if they did any of the following activities: interacted with friends; played mahjong, chess, or cards, or visited a community club; supported family, friends, or neighbors; went to a sport, social, or other kind of club; participated in a community-related organization; did volunteer or charity work; cared for a sick or disabled adult; attended an educational or training course; invested in stocks; surfed the Internet; and others. Health behaviors include smoking, alcohol drinking status, sleep duration, and number of chronic diseases. Smoking status was categorized into never smoker, ever smoker, and current smoker. Alcohol drinking status was categorized into never drinker and ever drinker. Chronic diseases included hypertension, diabetes or high blood sugar, cancer or malignant tumor, chronic lung disease, heart problems, stroke, kidney disease, stomach or other digestive diseases, arthritis or rheumatism, and asthma. Number of chronic diseases was categorized into 0, 1, 2, and ≥ 3.

### 2.5. Statistical analyses

#### 2.5.1 Descriptive statistics

Distributions of the baseline characteristics were described by stratifying household solid fuel. Means ± standard deviations (SDs) and numbers (percentages) were used to describe the continuous and categorical variables, respectively. Mann-Whitney U test and Chi-square test were used to examine the differences in continuous variables and categorical variables, respectively.

#### 2.5.2 Single exposure models

Generalized estimating equations (GEEs) were used to separately examine the associations of solid fuel use for cooking or heating with BDRM scores. In Model 1, we adjusted for sex. In Model 2, we further adjusted for residence, marital status, participation in social activities, alcohol drinking status, smoking status, sleep duration, number of chronic diseases, and waves. We estimated the coefficients and their 95% confidence intervals (CIs) for cooking or heating in the two GEE models.

#### 2.5.3 Co-exposure models

In order to further compare the contributions of cooking and heating, we examined the joint associations of solid fuel use for cooking and heating with BDRM scores by adding two fuel types and their interaction in GEE models. Since no significant interaction was observed, we removed it in the final models. Covariates in two GEE models were the same as those in the single-exposure models. We reported the coefficients and 95% CIs for cooking and heating in GEE models.

Additionally, we investigated the associations of BDRM score with the following four exposure groups: group 1 (clean cooking fuel type & clean heating fuel type), group 2 (solid cooking fuel type & clean heating fuel type), group 3 (clean cooking fuel type & solid heating fuel type) and group 4 (solid cooking fuel type & solid heating fuel type). Two GEE models using the above four groups as the exposure variable were performed, adjusting for the same covariates as mentioned above. We reported the coefficients and their 95% CIs for the four exposure groups in the two GEE models.

#### 2.5.4 Subgroup analyses

To examine whether the associations of solid fuel use for cooking and heating with BDRM scores differed by subgroup, we stratified the single-exposure models by baseline characteristics including age (≤ 65 years, > 65 years), sex (male, female), residence (rural, urban), marital status (live with spouse, live without spouse), participation in social activities (yes, not), alcohol drinking status (never drinker, ever drinker), smoking status (never smoker, ever smoker, current smoker), sleep duration (> 4 and < 10 hours per night, ≤ 4 or ≥ 10 hours per night) (Ma et al. 2020), and number of chronic disease (0-1, ≥ 2). We estimated the coefficients and their 95% CIs for cooking or heating.

All statistical analyses were performed using R 4.1.3 (http://www.R-project.org). The “geepack” packages were used for the GEE model (Hjsgaard et al. 2005). A two-sided P value of <0.05 was considered statistically significant.

#### 2.5.5 Sensitivity analyses

To test the robustness of our findings, we conducted the sensitivity analyses including: (1) excluding participants with extremely high (i.e., > mean + 2SD) BDRM scores at baseline; (2) using another dementia risk prediction model, BDSI (Brief Dementia Screening Indicator)(Barnes et al. 2014); (3) further adjusting for more covariates in Model 2.

## 3. Results

### 3.1. Baseline characteristics of the study population

Table1 shows the baseline characteristics of participants included in the prospective analysis. Participants who used solid (vs. clean) fuel tended to be older (e.g., for cooking, 59.91 vs 58.15 years), live in rural areas (e.g., for cooking, 77.9% vs 37.9%), lessly participate in social activities (e.g., for cooking, 44.7% vs 56.3%), and have more chronic diseases (e.g., for cooking, 1.36 vs 1.21). Moreover, participants who used solid (vs. clean) fuel had a significantly higher BDRM score.

### 3.2. Association of solid fuel use for cooking and heating with BDRM scores

Figure 2 presents the associations of solid fuel use for cooking and heating with BDRM scores from single-exposure models. Participants who used solid (vs. clean) fuel had significantly higher BDRM scores in all models. For instance, in fully adjusted Model 2, participants who used solid (vs. clean) fuel for cooking or heating had a 0.14 (95% CI: 0.12, 0.17, P < 0.001) and 0.11 (95% CI: 0.08, 0.13, P < 0.001) increase in BDRM scores, respectively.

**Fig. 2.**
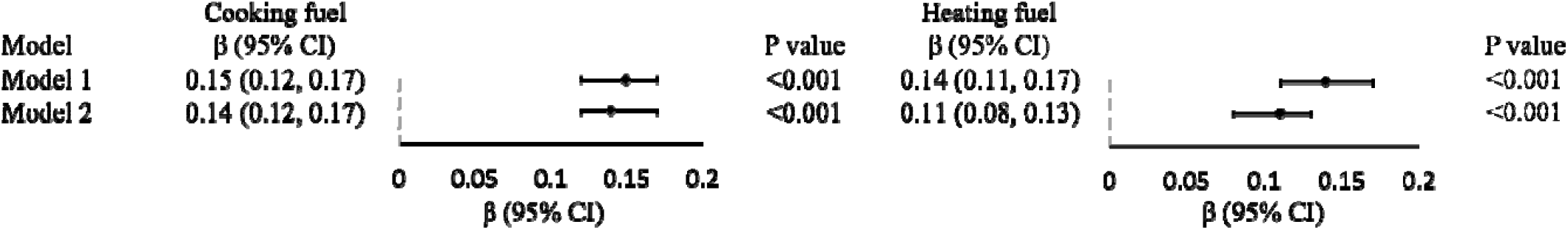
Associations of solid fuel use for cooking and heating with BDRM scores. Note: BDRM, Rotterdam Study Basic Dementia Risk Model; ß, regression coefficient; Cl, confidence interval.

Model 1 adjusted for sex. Model 2 adjusted for sex, residence, marital status, participation in social activities, alcohol drinking status, smoking status, sleep duration, number of chronic diseases and wave.

### 3.3 Joint association of solid fuel use for cooking and heating with BDRM scores

Table 2 shows the joint associations of solid fuel use for cooking and heating with BDRM scores from co-exposure models. We found that solid fuel use for both cooking and heating was independently associated with BDRM scores. For instance, in fully adjusted Model 2, participants who used solid fuel for cooking had a 0.16 (95% CI: 0.13, 0.19, P < 0.001) increase in BDRM scores and participants who used solid (vs. clean) fuel for heating had a 0.03 (95%CI: 0.00, 0.06, P = 0.040) increase in BDRM scores. Meanwhile, group 4 (solid cooking fuel & solid heating fuel) had the highest BDRM score,with a ß of 0.20 (95% CI: 0.16, 0.23, P < 0.001), relative to group 1 (clean cooking fuel type & clean heating fuel type).

**Table 1.**
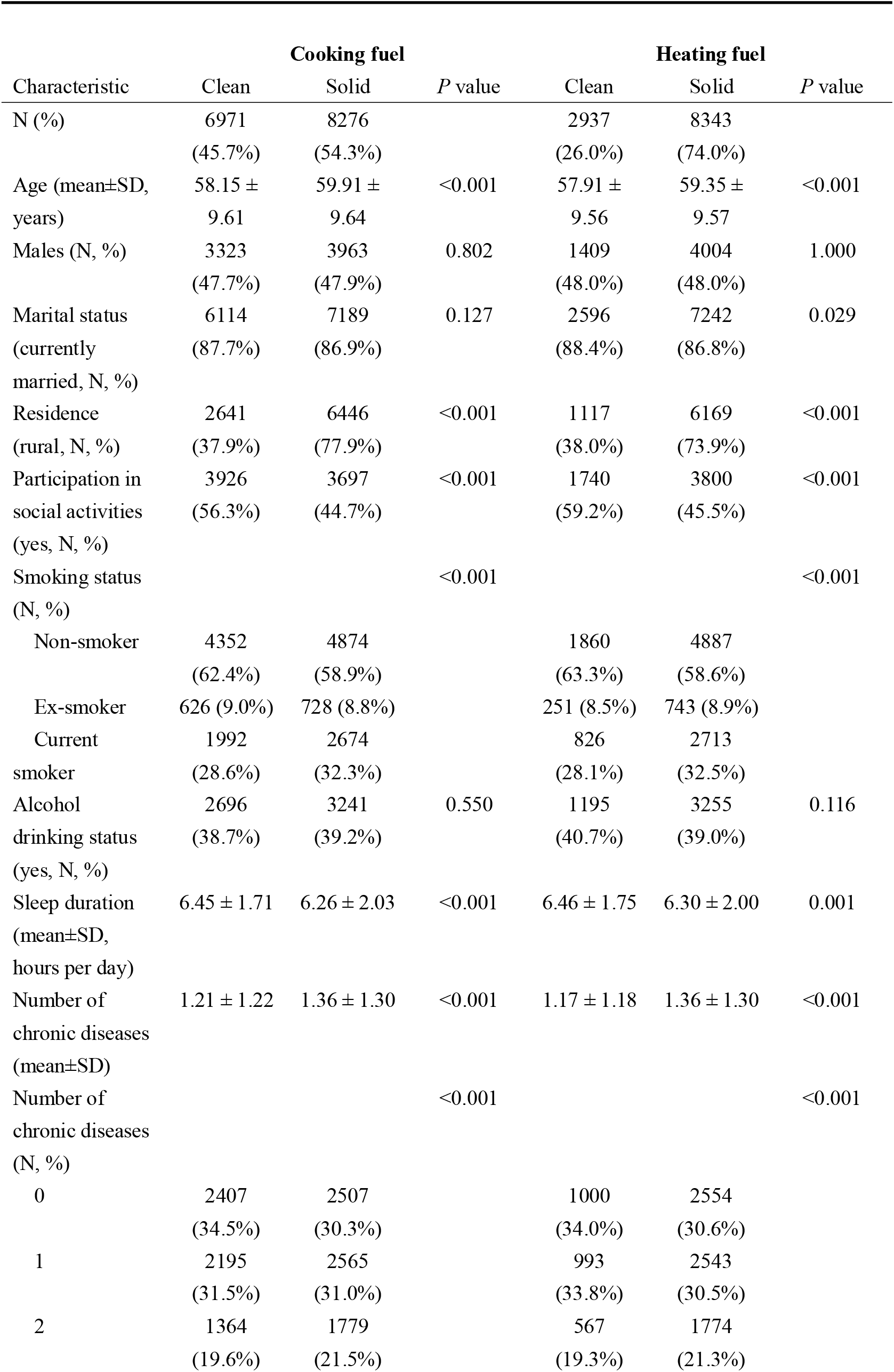
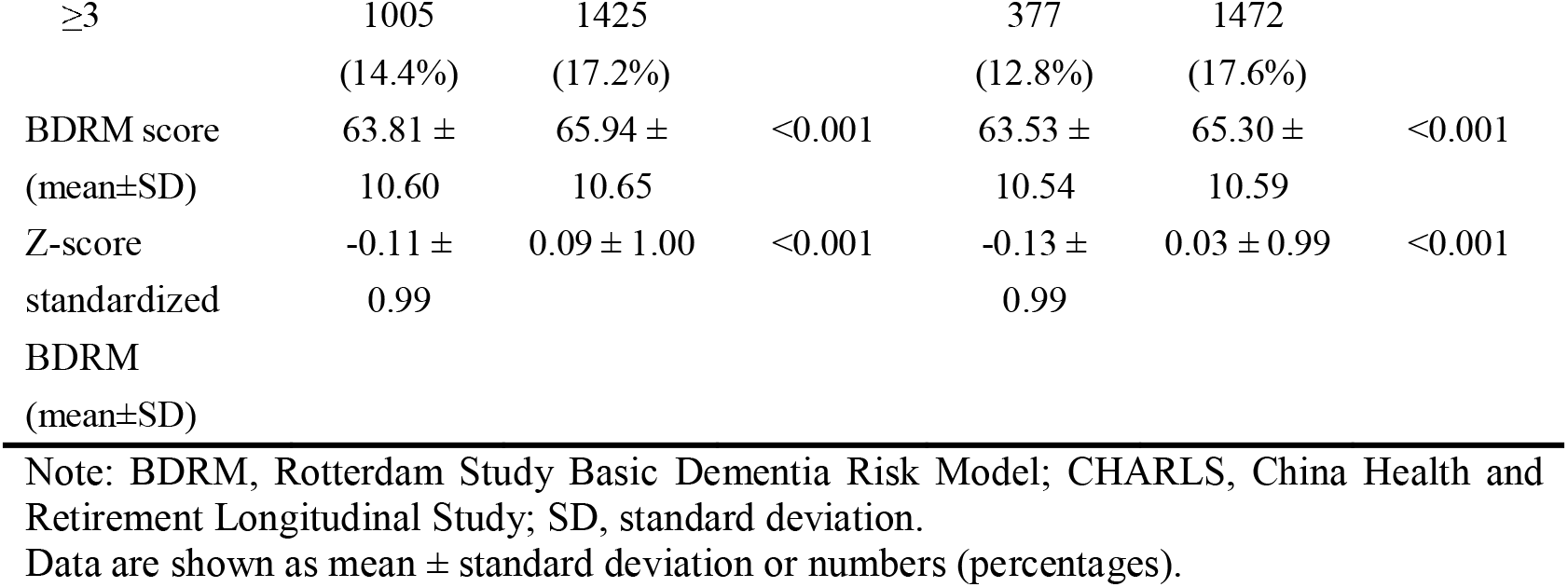
The baseline characteristics of participants by types of household fuels in CHARLS 2011.

**Table 2.** Joint associations of solid fuel use for cooking and heating with BDRM scores.

| | $\beta$ (95% CI ) | *P* value |
| --- | --- | --- |
| **Exposure of solid cooking and solid heating** |  |  |
| **Model 1** |  |  |
| Cooking | 0.17 (0.14, 0.20) | <0.001 |
| Heating | 0.05 (0.02, 0.08) | 0.003 |
| **Model 2** |  |  |
| Cooking | 0.16 (0.13, 0.19) | <0.001 |
| Heating | 0.03 (0.00, 0.06) | 0.040 |
| **Exposure of grouped clean/solid cooking and heating** |  |  |
| **Model 1** |  |  |
| Group1: clean cooking fuel & clean heating fuel (reference) |  |  |
| Group2: solid cooking fuel & clean heating fuel | 0.18 (0.12, 0.25) | <0.001 |
| Group3: clean cooking fuel & solid heating fuel | 0.05 (0.02, 0.09) | 0.007 |
| Group4: solid cooking fuel & solid heating fuel | 0.22 (0.19, 0.25) | <0.001 |
| **Model 2** |  |  |
| Group1: clean cooking fuel & clean heating fuel (reference) |  |  |
| Group2: solid cooking fuel & clean heating fuel | 0.20 (0.14, 0.26) | <0.001 |
| Group3: clean cooking fuel & solid heating fuel | 0.05 (0.01, 0.09) | 0.010 |
| Group4: solid cooking fuel & solid heating fuel | 0.20 (0.16, 0.23) | <0.001 |

Note: BDRM, Rotterdam Study Basic Dementia Risk Model; $\beta$ , regression coefficient; CI, confidence interval. Model 1 adjusted for sex. Model 2 adjusted for sex, residence, marital status, participation in social activities, alcohol drinking status, smoking status, sleep duration, number of chronic diseases and wave.

### 3.4 Association of solid fuel use for cooking and heating with BDRM scores by subgroups

The results of subgroup analyses are shown in Figures 3 and 4, respectively. Overall, the associations of solid fuel use for cooking and heating with BDRM scores were largely maintained in different subgroups. We observed the stronger associations in participants living in rural areas and without comorbidity. For instance, in rural areas, participants who used solid (vs. clean) fuel for cooking had a 0.20 (95% CI: 0.17, 0.23) increase in BDRM score and in urban areas, the estimate was 0.06 (95% CI: 0.02, 0.11).

**Fig. 3.**
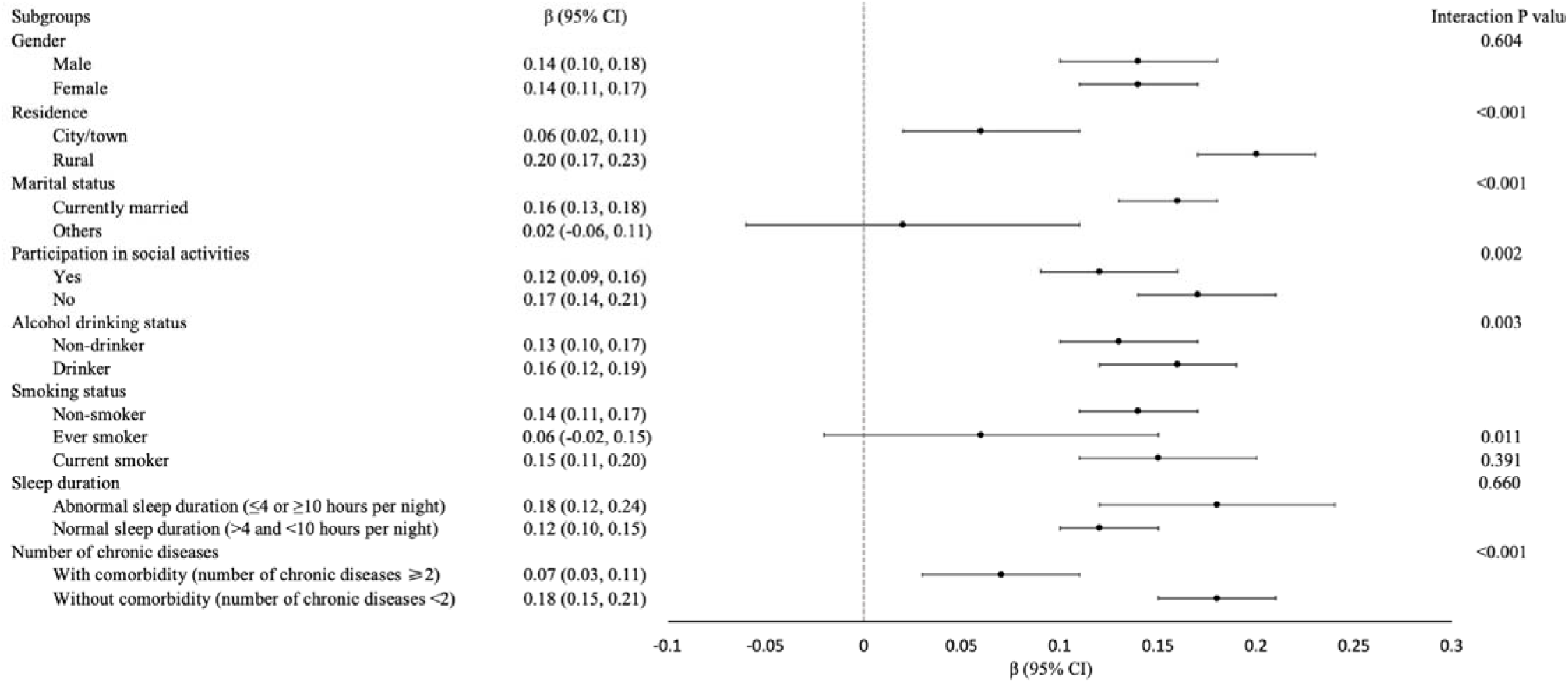
Subgroup analyses of associations of solid fuel use for cooking with BDRM scores. Note: BDRM, Rotterdam Study Basic Dementia Risk Model; ß, regression coefficient; Cl, confidence interval.

**Fig. 4.**
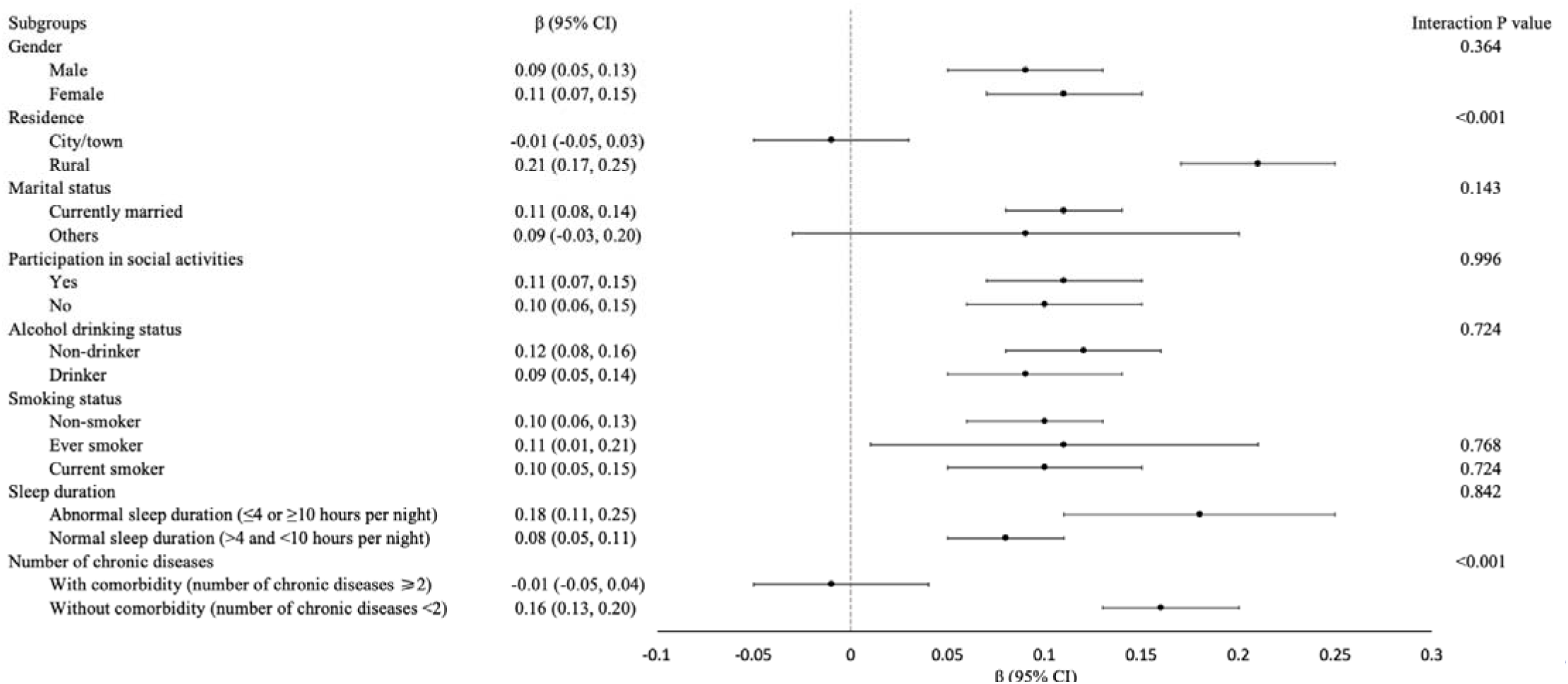
Subgroup analyses of associations of solid fuel use for heating with BDRM scores. Note: Data: ß (95% CI), Interaction p-value;outcome variable: dementia risk scores. BDRM, Rotterdam Study Basic Dementia Risk Model; ß, regression coefficient; Cl, confidence interval.

### 3.5. Sensitive analyses

First, excluding participants with an extremely high (>mean + 2SD) BDRM score at baseline did not alter the significant associations between solid fuel use and increased BDRM scores in all models (Table S1). Second, we found similar results using another dementia risk prediction model-BDSI (Table S2-S5 and Figure S1-S4).

For instance, as shown in Figure S2, in fully adjusted Model 2, participants who used solid fuel for cooking had a 0.23 (95% CI: 0.19, 0.28, P < 0.001) increase in BDSI scores and participants who used solid fuel for heating had a 0.16 (95% CI: 0.10, 0.21, P < 0.001) increase in BDSI scores. Third, the results were consistent with those from primary analyses after adjusting more covariates in Model 2 for BDSI and BDRM analysis, respectively (Table S7).

## 4. Discussion

Based on a large sample of middle-aged and older adults from a nationally representative cohort study, we found that solid fuel use for cooking and heating was independently associated with higher BDRM scores (i.e., increased dementia risk), and solid fuel use for cooking exhibited a greater effect on BDRM scores. The associations were maintained in most subgroups and stronger in participants living in rural areas. Our findings highlight the potential of using clean fuel indoors, especially for cooking, for preventing dementia in rural areas.

Nearly three billion people use solid fuels worldwide which is the main reason for HAP (Lim et al. 2012). In China, about one million people died prematurely because of the solid fuel used indoors (Lertxundi et al. 2015). Meanwhile, China is experiencing the highest growing number of people with dementia (Prince 2016). So far, several studies have examined the association of solid fuel use with negative health outcomes, including asthma (Thacher et al. 2013), lower respiratory tract infections (Ezzati and Kammen 2001), chronic obstructive pulmonary disease, all-cause mortality (Smith and Mehta 2003) and preeclampsia/eclampsia (Agrawal and Yamamoto 2015). However, no studies have explored the association of solid fuel use with dementia risk. For the first time, we demonstrated that solid fuel use for cooking and heating was associated with increased dementia risk. Our findings highlight an intriguing new avenue for dementia prevention and control, and calls for more efforts to facilitate universal access to clean energy, especially in LMICs such as China. In the Fourteenth Five Year Plan of Agriculture and Rural Area Modernization, China is seeking for evidence to initiate a series of policies to promote the utilization of clean energy, such as solar power and wind energy, and extend the construction of clean heating facilities. Further studies would be applied to explore the improvement of the cognitive function of people through the intervention of this clean energy revolution in China.

Our results in subgroup analysis showed a stronger association of solid (vs. clean) fuel use with increased dementia risk in participants who lived in rural areas. In China, adults in rural areas are still heavily reliant on solid fuel (Chan et al. 2017). Moreover, the prevalence of dementia in rural areas is higher than that in urban China (Hu et al., 2021) (Hu et al. 2021). Adults in rural areas might be more vulnerable to air pollutants caused by solid fuel use, due to their relatively poor health in comparison to their counterparts in urban (Zhang 2017). Nonetheless, an increasing number of Chinese people are willing to retire to the countryside, highlighting a critical intervention point to raise awareness among this group about the benefits of using clean fuel rather than solid fuel (Shen et al. 2021). Considering fewer dementia-related medicare resources in rural areas, this could provide a useful approach for preventing dementia. We also found a stronger association of solid (vs. clean) fuel use with a higher BDSI score in females. This was consistent with Chen’s study where solid fuel use was stronger associated with cognitive function decline among females than males in China (Chen et al. 2021). The plausible explanation is that females in China experience more exposure to HAP from solid fuel use because they actively participate in domestic activities such as cooking. Another possible explanation is that there was a sex difference in the biological effect of solid fuel use on human health, given that PM2.5 has a greater adverse effect in females than in males (Eze et al. 2015; Kim et al. 2019). The above findings may aid in the identification of vulnerable subgroups such as those living in rural areas or females, and the development of precise prevention programs for dementia.

The underlying biological mechanisms of the association observed are unclear. Solid fuel use produces high levels of gaseous pollutants (e.g., CO, NOx, O_3_) and particulate matters (e.g., PM2.5) (Chen et al. 2016; Clark et al. 2013; Ni et al. 2016). The particles could translocate to the brain through the circulation crossing the blood-brain barrier (Block and Calderón-Garcidueñas 2009) or olfactory neurons (Tonelli and Postolache 2010) and then cause neurotoxicity (Terzano et al. 2010). The particles trigger dementia pathogenesis via multiple pathways (Akiyama et al. 2000; Block et al. 2012; Calderón-Garcidueñas et al. 2002; Costa et al. 2014). First, particles contribute to dementia pathogenesis by inducing neuroinflammation and oxidative stress (Peters et al. 2006). The second plausible pathway involves the promotion or aggravation of cardiovascular effects. PM2.5 and other pollutants (e.g., NO_2_) have been demonstrated to have adverse effects on cardiovascular health, a well-known risk factor for cognitive decline and dementia (O’Brien 2006; Stampfer 2006). Vascular disease may have an indirect effect, i.e., vascular injury in the brain may produce conditions that predispose to neurodegeneration. Alternatively, vascular factors may play a direct role in Alzheimer’s disease (AD) progression by promoting neuronal death and the formation of plaques and tangles (the hallmark pathology of Alzheimer’s disease) (Stampfer 2006). Further studies are needed to explore the toxicological mechanisms of solid fuel use with dementia risk.

Our study has some strengths. First, this study conducted a comprehensive analysis of the association between the use of solid fuel and dementia risk, using a prospective cohort design and a relatively large sample size of middle-aged and older adults. Second, we used two widely-used dementia risk models of BDSI and BDRM, and the consistent results showed the robustness of our findings. Third, using dementia risk as an outcome enables us to track and prevent dementia at an earlier stage. However, the study has several limitations. First, data on solid fuel use was self-reported; and thus, recall bias is possible. Second, data on detailed exposure duration for solid fuel use were unavailable; and thus, we could not evaluate the effect of exposure duration on dementia risk. Third, outdoor air pollution was not controlled, and it may bias the results. Fourth, BDSI and BDRM are not developed originally in China; however, the two models have been proved to show well discriminative ability when used in LMICs (Stephan et al. 2020).

## 5. Conclusions

For the first time, this study demonstrated that solid fuel use for cooking and heating was independently associated with increased dementia risk among Chinese middle-aged and older adults, particularly among those living in rural areas. Our findings call for more efforts to facilitate universal access to clean energy for dementia prevention. Meanwhile, more studies are needed to disentangle the toxicological mechanisms of solid fuel use with dementia risk.

## Data Availability

All data produced are available online at http://charls.pku.edu.cn

## Abbreviations

*L*MICs: low- and middle-income countries
HICs: high-income countries
HAP: household air pollution
BDRM: Rotterdam Study Basic Dementia Risk Model
CHARLS: China Health and Retirement Longitudinal Study
BDSI: Brief Dementia Screening Indicator
SD: standard deviation
CI: confidence interval
CVD: cardiovascular disease
AD: Alzheimer’s disease
GEE: generalized estimating equations

## Acknowledgments

This research was supported by a grant from a grant from the National Natural Science Foundation of China (72004201), the Fundamental Research Funds for the Central Universities (ZL), Key Laboratory of Intelligent Preventive Medicine of Zhejiang Province (2020E10004), Zhejiang University Global Partnership Fund (188170-11103), and a grant of Humanity and Social Science Youth Foundation of Ministry of Education (20YJC840019). The funders had no role in the study design; data collection, analysis, or interpretation; in the writing of the report; or in the decision to submit the article for publication.

